# Sexually transmitted infections among key populations in India: systematic review with spatiotemporal distribution

**DOI:** 10.1101/2024.05.06.24306926

**Authors:** Mihir Bhatta, Agniva Majumdar, Subrata Biswas, Utsha Ghosh, Piyali Ghosh, Papiya Banerji, Santhakumar Aridoss, Bhumika Tumkur Venkatesh, Nibedita Das, Abhisek Royal, Protim Ray, Prabuddha Gopal Goswami, Turalapati Laxmi Narayan Prasad, Gajendra Kumar Medhi, Pankaj Kumar Khan, Lahari Saikia, Falguni Debnath, Debjit Chakraborty, Alok Kumar Deb, Rajatsuvra Adhikary, Shanta Dutta

## Abstract

In the developing world, sexually transmitted infections (STIs) are among the key sources of health and financial adversities, contributing significantly to morbidity, death, and stigma. The frequency and prevalence of four treatable STIs— syphilis, chlamydia, gonorrhea and trichomoniasis— vary greatly throughout different geographical regions. In the current situation, published reports from multiple local, national and international bodies are combined with information from peer-reviewed published articles to generate present systematic review. Goal of the current systematic review is to identify the geographic distribution and current of STIs among the Indian key population. The PRISMA flow diagram describes the specific criteria of inclusion and exclusion as well as the quantity of articles in each category. A relevant electronic database search produced to find 40 publications that matched the inclusion criteria outlined in the protocol for the current systematic review were found after a search of pertinent electronic databases However, it was found that published articles were not available from all of the geographical regions in India. The majority of the research was found in the western and southern regions. Few studies from the northern and north eastern regions of India have been published. The majority of published studies on STI prevalence were based on syphilis seroprevalence and focused on the MSM (men sex with men) and FSW (female sex workers) populations, followed by H/TG (Hijras with Transgenders) and PWID (those who inject drugs). The majority of the research used aetiological diagnosis to report prevalence. It can be concluded in light of the current findings with considering the noted limitations, present HIV surveillance system under the NACP (National AIDS Control Programme) may utilized the collected extra bio-specimen to establish prevalence of STIs in high risk populations. However, in coming days, with the availability of the comparable data from most of the regions in India, it will be possible to perform a systemic review and meta-analysis on the spatiotemporal distribution of the four curable STIs in Indian general population.

## Introduction

In the developing world, sexually transmitted infections (STIs) are among the key sources of health, and financial adversities, causing extensive morbidity, mortality and stigma [1]. A remarkable variation of frequency and prevalence of the four curable STIs, which are syphilis, chlamydia, gonorrhoea and trichomoniasis are observed with the spatiotemporal variation [2]. Studies suggest that the prevalence of these four STIs among general people has a range of 0-3.9% in India [3], but the STI burden is considerably higher among key populations having high-risk behaviour like men who have sex with men (MSM), female sex worker (FSW) hijra/transgender, and people who inject drugs (PWID). However, there is inadequate literature to date reported STI burden in Indian key populations and all of the information have not been unified to enlighten the general spatiotemporal tendencies of STI infections in various key or high-risk population. Present article was set with this intention to extract information on prevalence of four curable sexually transmitted infections viz. syphilis (by *Treponema pallidum*), gonorrhoea(by *Neisseria gonorrhoeae* or NG), chlamydia (by *Chlamydia trachomatis* or CT), and trichomoniasis (by *Trichomonas vaginalis* or TV) among four key populations such are, men who have sex with men (MSM), female sex workers (FSW), hijras with transgenders (H/TG) and people who inject drugs (PWID) in India and to perform a systematic review of available evidence on the prevalence and the spatiotemporal distribution of these four STIs in Indian key population [3–4].

To gain insight of the status of the prevalence and spatiotemporal distribution of STIs among key population in India, present systematic review has been initiated using existing previously published evidence. The present findings would be vital to understand the STI status among key population and to design evidence-based strategies for STI prevention in key population in India.

## Materials and methods

A study protocol is developed according to the Preferred Reporting Items for Systematic Reviews and Meta-Analysis (PRISMA) guidelines. All available peer reviewed published articles are extracted using suitable search terms in PUBMED, MEDLINE, EMBASE, Cochrane Library, Science Direct, Google Scholar and Psychinfo along with published reports (as grey literature) from reliable sources, comprising all studies from January 2000 to December 2023.

A protocol for the systematic review was prepared and registered in the PROSPERO, having the registration number CRD42022357425 and published subsequently [5].

### Evaluation of the methodological quality

Evaluation of selected studies was performed through title, text and abstract prior to the accumulation of it into the decisive analysis. Evaluation was performed with the help of modified Newcastle - Ottawa Quality Assessment tool [6]. Evaluation was completed before collection of information.

### Geographical distribution with spatiotemporal attributes

Geographical distributions of previously mentioned STIs in Indian key population were presented using heat map and choropleth maps, generated with the help of *Quantum GIS (Ver. 3.18 Zurich)* [7]. An administrative map of India containing the latest information of states and union territories is georeferenced and the resulting polygon was used as the base map layer. State-level STI prevalence data were added as comma delimited layers. Prevalence data from accepted studies were added as spatiotemporal attributes in separate layers respectively. Finally, maps were generated for overall included studies for each key population with prevalence of four curable STIs in histogram format in a separate layer. Multiple points were used to generate heat map of STI prevalence. Data based on regional distribution of STIs among key population has been entered into the newly prepared maps as non-spatial attributes [8].

## Results

There are total of 8919 publications are selected initially. The abstract and titles were screened for eligibility and duplicates were removed. A total of forty articles were included from peer reviewed journals. Among the 70 reports (grey literature)-excluding Sankalak Reports (two documents), published by several competent national, international, and state agencies, any reports could not be included based on the screening tool. However, observations from four reports were presented due to availability of few relevant data. Details of the inclusion and exclusion criteria, along with the, number of each category of articles are described through PRISMA flow Diagram [Figure 1]. Search of relevant databases (PUBMED. Google Scholar, Mendeley, Cochrane Library, Scopus, Science Direct and EMBASE) yielded 40 articles which met the inclusion criteria set in the protocol for the present systematic review. However, it is found that published articles were not available from all geographical regions in India. Most of the studies were conducted in southern and western part. However, four studies were also reported from north-eastern part of India. A few studies form northern India was also conducted in Delhi and Lucknow [Figure 2].

**Figure 1.**
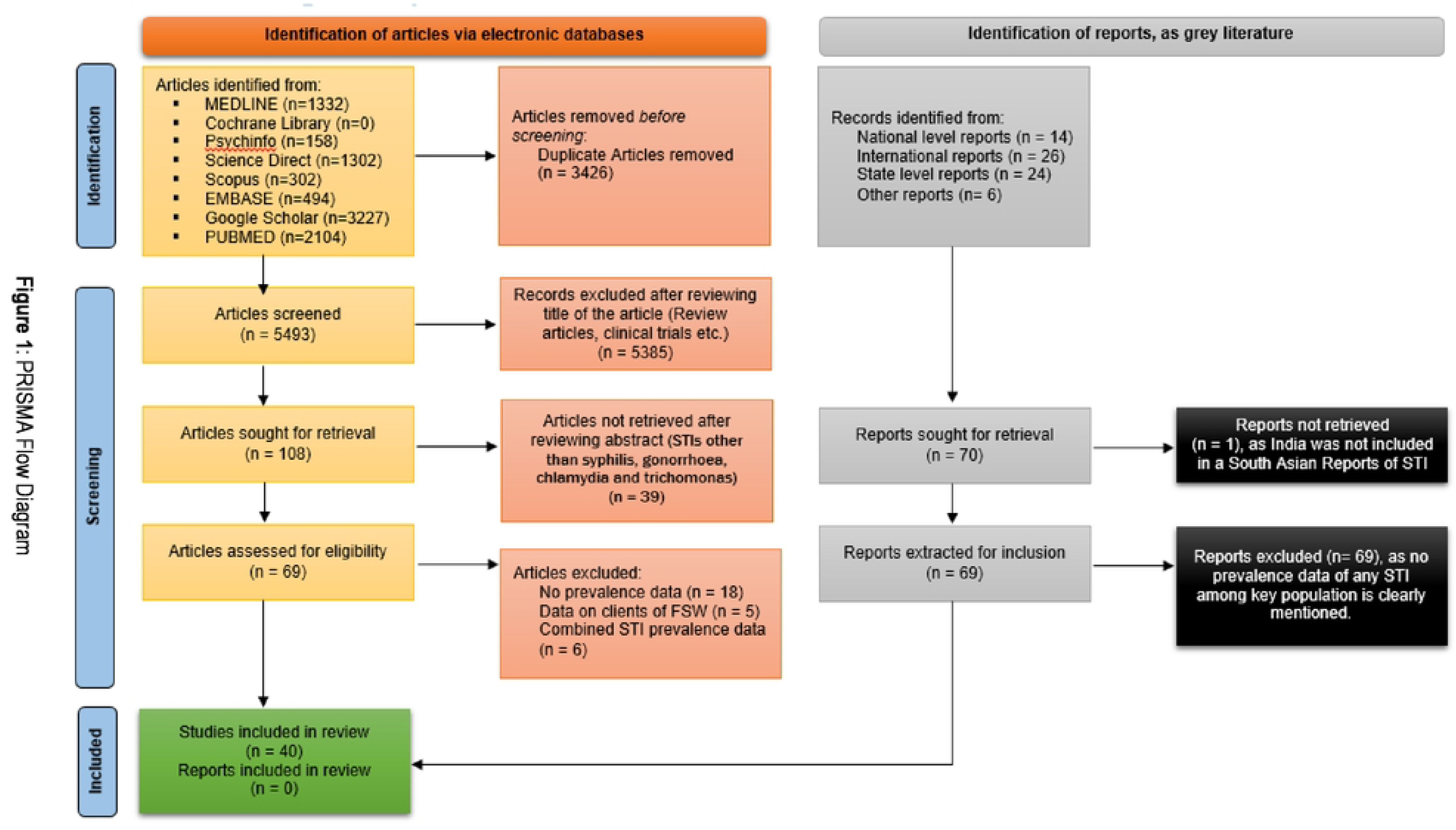
PRISMA Flow Diagram.

**Figure 2:**
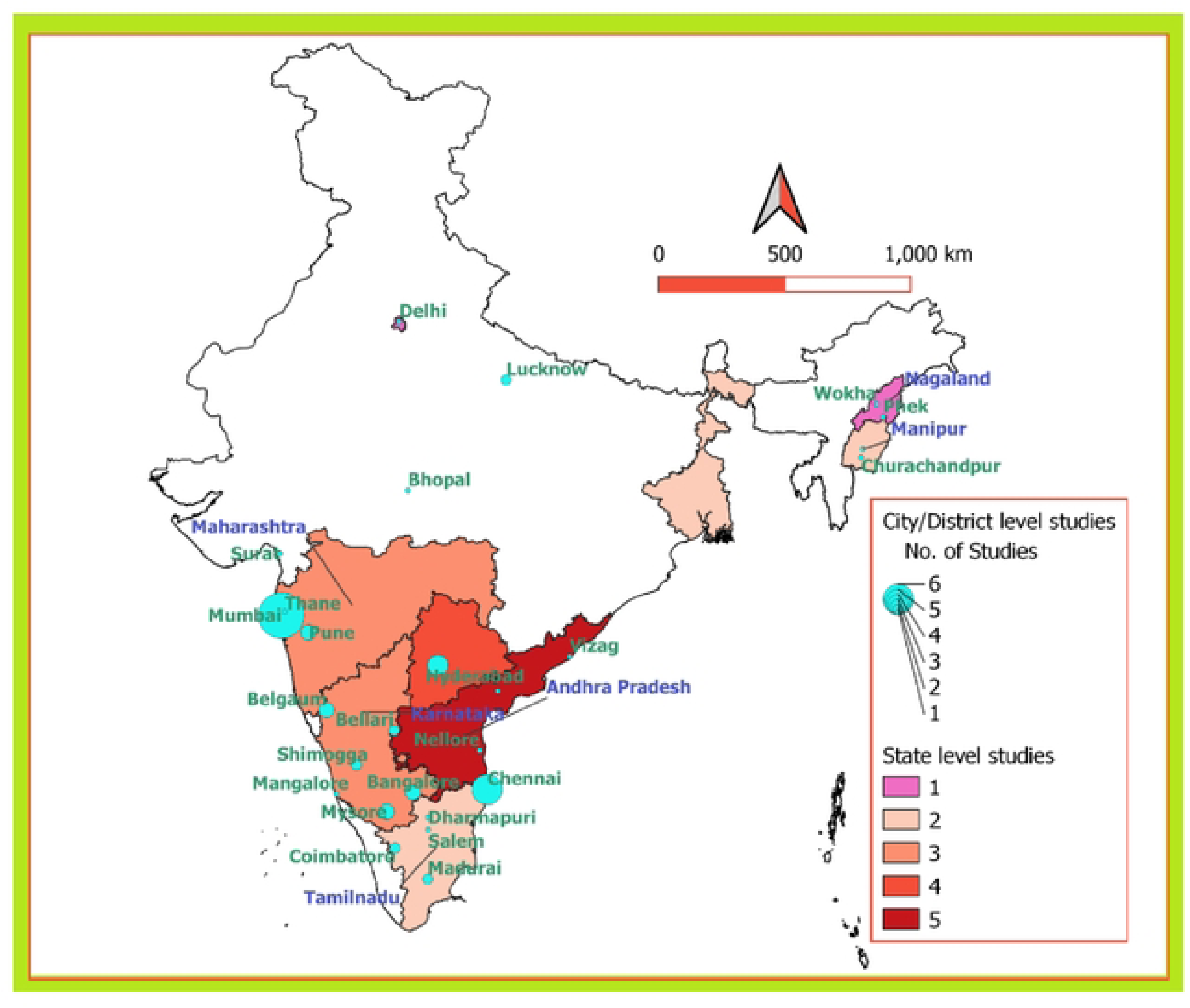
Map of India indicating the regions where included studies were conducted.

Maximum numbers of reviewed studies reported on STI prevalence were among MSM and FSW population groups followed by H/TG and PWID and mostly based on syphilis seroprevalence. Data on chlamydia and gonorrhoea were also available but much less in number, trichomoniasis was least reported STI. While synthesizing data, articles which included more than one key population was counted separately. Eighteen articles reported STI prevalence among FSWs [1–4, 8–23]. These eighteen articles included twenty-six different studies based on prevalence data of different STI, FSW subgroups and geographical region. All data related to FSW was presented under two different diagnostic methods, aetiological [1–4, 8–20] [Table 1] and syndromic [20,23] [Table 2]. No study was reported after 2021. STI prevalence among MSM was reported in nineteen articles [3,15, 24–40]. Based on different STI and geographical region total forty studies were recorded for this review. Relevant extracted data on MSM was recorded under aetiological diagnosis [3,15, 24–40] [Table 3] and syndromic [33, 40] [Table 4] diagnosis. Four articles on H/TG included four studies on STI prevalence among H/TG based on different STI and geographical region [18, 29, 31, 41]. Data on H/TG under aetiological diagnosis was presented in Table 5. There was no data available for syndromic diagnosis. Two articles [15, 42] were available for STI prevalence among PWID. These two articles comprise six studies on prevalence of different STIs among PWID population in India [Table 6].

### Findings among FSW

The four curable STIs among FSW were examined in eighteen articles [Table 1 and 2], most of these conducted in Andhra Pradesh, Telangana, Karnataka, Maharashtra, Uttar Pradesh, West Bengal, and Nagaland. Almost half of the studies were conducted in facilities like STI clinics or tertiary care hospitals [Figure 3]. Out of 26 studies, most of the studies reported prevalence through aetiological diagnosis [Table 1]. Among the published articles during study period, article from Beksinska et al. was published in 2018 [22], however, the study period was 2011. There was any published article on STI prevalence estimation study among FSW in India that was conducted after 2011. It was a cross-sectional study on syphilis among FSW in Karnataka and the estimated prevalence was 3.2. Screening was done through RPR followed by TPHA. Most of the studies reported on single STI, namely syphilis. Das et al. [17] reported on all four STIs among FSW population. This facility level cohort study was conducted in Mumbai and Hyderabad. Diagnosis of syphilis was done through RPR followed by TPHA from blood sample. Estimated prevalence of Syphilis was 5.8. Chlamydia, Gonorrhoea, and Trichomoniasis diagnosis were done through Nucleic Acid Amplification Test (NAAT) and reported prevalence were 16.1, 14.2 and 31.2 respectively. Out of all reviewed studies on FSW, the study by Ghosh et al [15] estimated very high syphilis prevalence (28.8) among FSW in West-Bengal. This is also a facility level cross-sectional study with sample size of 45. Gupte et al. [11] published research article on different subgroups of FSW in 2011. This cohort study was conducted in Mumbai during the period of 2007-2008. Prevalence of syphilis was estimated in this study. Brothel based FSW was with highest syphilis prevalence of 6.6 and the lowest prevalence (1.3) was reported by bar based FSWs. Trichomoniasis was the least examined and hence least reported STI among FSW [Figure. 3]. Syndromic diagnosis unveiled genital herpetic and non-herpetic ulcer, cervical discharge, Inguinal buboes vaginal discharge and ano-rectal discharge for FSW population [Table 2]. The highest reported symptom was vaginal discharge (50.7%) in FSWs. This was reported by Sarna et al [20]. This cohort study was conducted in Nellore, Andhra Pradesh during the period of January to December, 2011. There was a single state level entry each from Nagaland, West Bengal, and Maharashtra. Two state level entries from Karnataka and Tamil Nadu were also reported. Three state level entries from Andhra Pradesh were observed. Most of the district level studies were reported from Karnataka and Andhra Pradesh. Four studies from Mumbai and two each from Bangalore and Mysore were reported [Figure 3].

**Figure 3:**
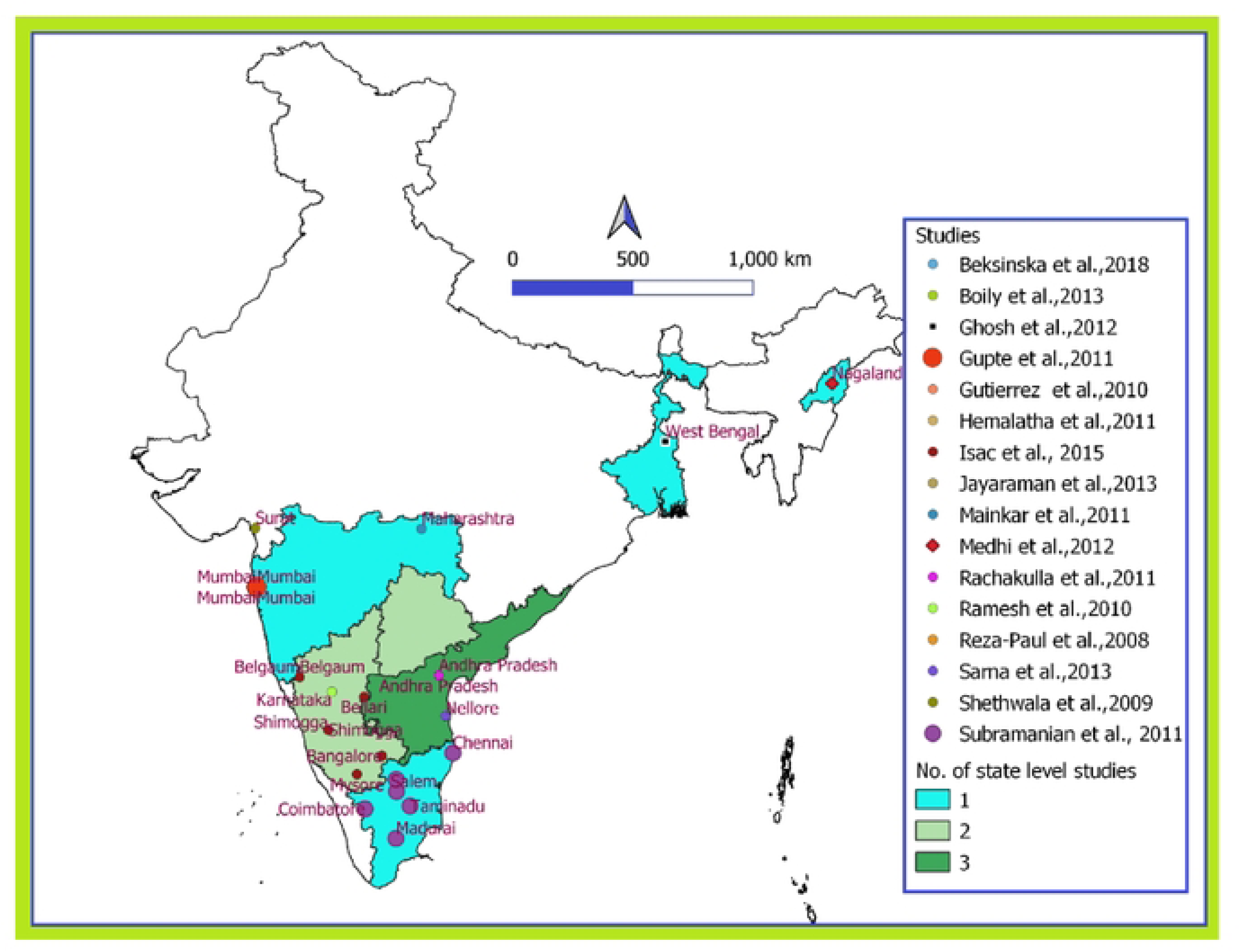
Representation of published articles on four curable STIs among FSWs in India.

### Findings among MSM

Nineteen articles [3, 15, 24–40] on MSM were included in the present study. Most of them were conducted in Tamil Nadu, West Bengal, Andhra Pradesh, Telangana, Karnataka, and Maharashtra [Figure 4]. Almost half of the studies reported from STI clinics or tertiary care hospitals. Most of the studies reported prevalence through aetiological diagnosis [Table 3].

**Figure 4:**
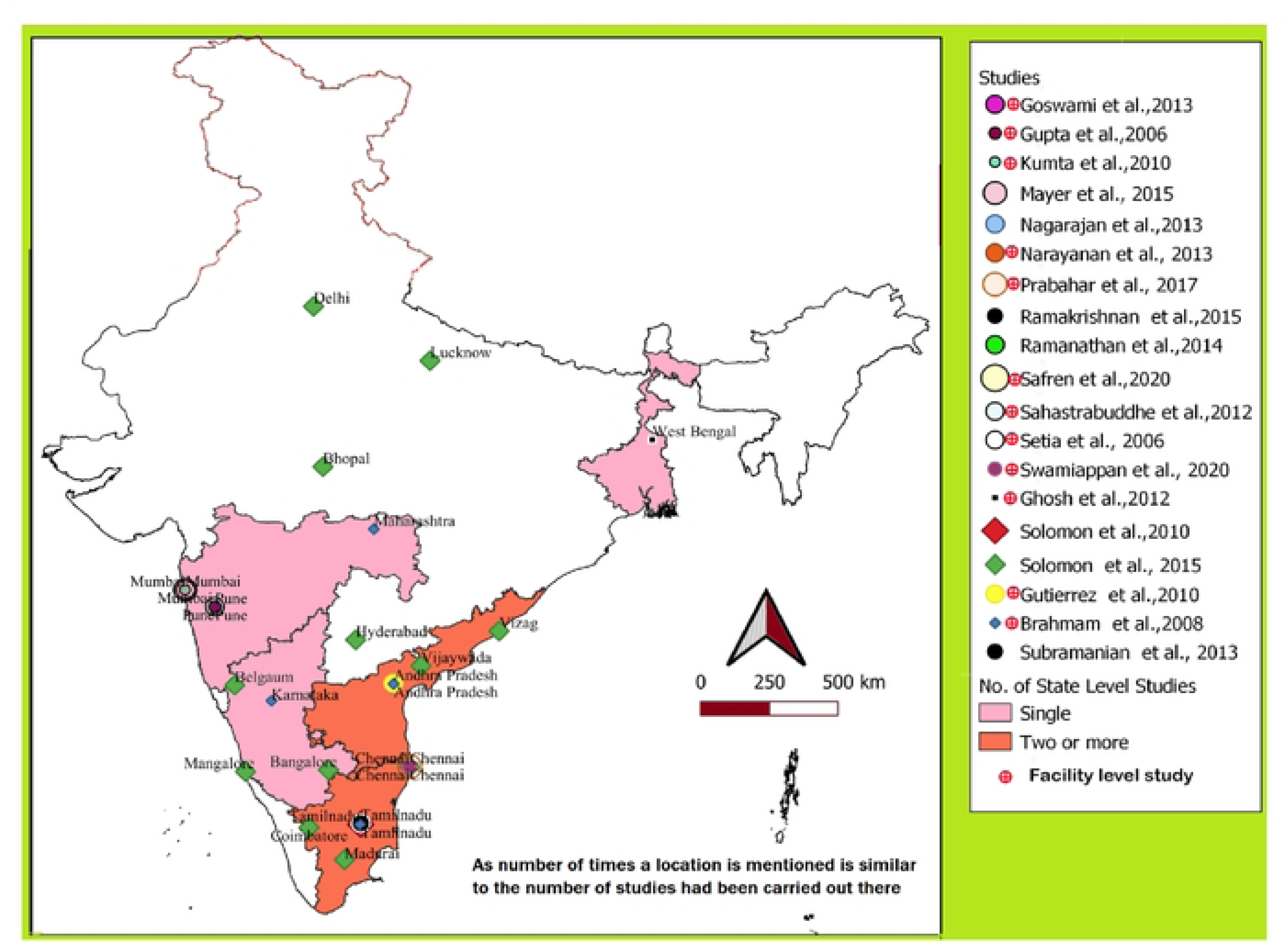
Representation of published articles on four curable STls among MSMs in India.

The most recent available publication for STIs among MSM was by Swamiappan et al. [40] and by Safren et al. [39] both was published in 2020. The cohort study was conducted in Chennai during July 2016 to June 2019 by Swamiappan et al., [40] and reported prevalence for syphilis for MSM. Syphilis screening was done through RPR without mentioning any confirmatory test with the prevalence of 8.8. Swamiappan et al. also reported symptoms of Urethral Discharge (2%), Genital herpetic Ulcer (3.1%) and Non-herpetic Ulcer (2%). Safren et al. [39] did their study in Mumbai and Chennai without specifying any study period [Table 4]. This cross-sectional study reported prevalence of syphilis, gonorrhoea, and chlamydia to be 16.1, 8.3 and 11.3 respectively in Chennai and 14.8, 16, 18.2 respectively in Mumbai. Most of the reviewed studies examined Syphilis. None of the studies reported Trichomoniasis among MSM. Gonorrhoea and chlamydia were also examined by many of these studies. For aetiological diagnosis RPR screening test followed by TPHA confirmatory test were administered for syphilis and PCR and NAAT for gonorrhoea and chlamydia in most of the studies. A cross-sectional community-based study among MSM people, published by Gutierrez et al. [3] in Andhra Pradesh, estimated a very high syphilis prevalence of 20.0, during the period of 2003 to 2007. The most recent study in Andhra Pradesh on MSM, published by Solomon et al. [28], was conducted in Vijayawada and Vizag. This cross-sectional study, conducted during 2012-2013 had estimated prevalence of was 4.4, and 2.1 respectively in Vijayawada and Vizag.

One from each state of Karnataka, West Bengal and Maharashtra was reported whereas; two state level studies were reported from Andhra Pradesh and Tamil Nadu. A considerable number of studies were reported from Mumbai, Chennai, and Pune. A few studies were also included from Delhi, Lucknow, Bhopal, and Hyderabad. There was no published article from North-eastern India. Studies from Mumbai, Chennai and Pune showed higher prevalence among MSM population. Similar to FSW population, prevalence of syphilis was reported in most of the studies, whereas a fewer study reported on trichomonas among MSM people in India [Figure 4].

### Findings among H/TG

Four studies [11, 29, 31, 41] were reported on H/ TG population during study timeline [Figure 5]. The most recently published study was Dasarathan and Kalaivani [41]. This facility level cohort study was conducted in Chennai city during 2011 to 2014 with a sub group of males to female. The prevalence of syphilis was estimated at 20.7, for this study [Figure 5]. Only a single state level report was published from Tamil Nadu. Only three district level reports were published from Mumbai, Chennai, and Pune. Only syphilis data was reported in these studies. It showed a poor regional representation of STI prevalence in India for H/TG population [Table 5].

**Figure 5:**
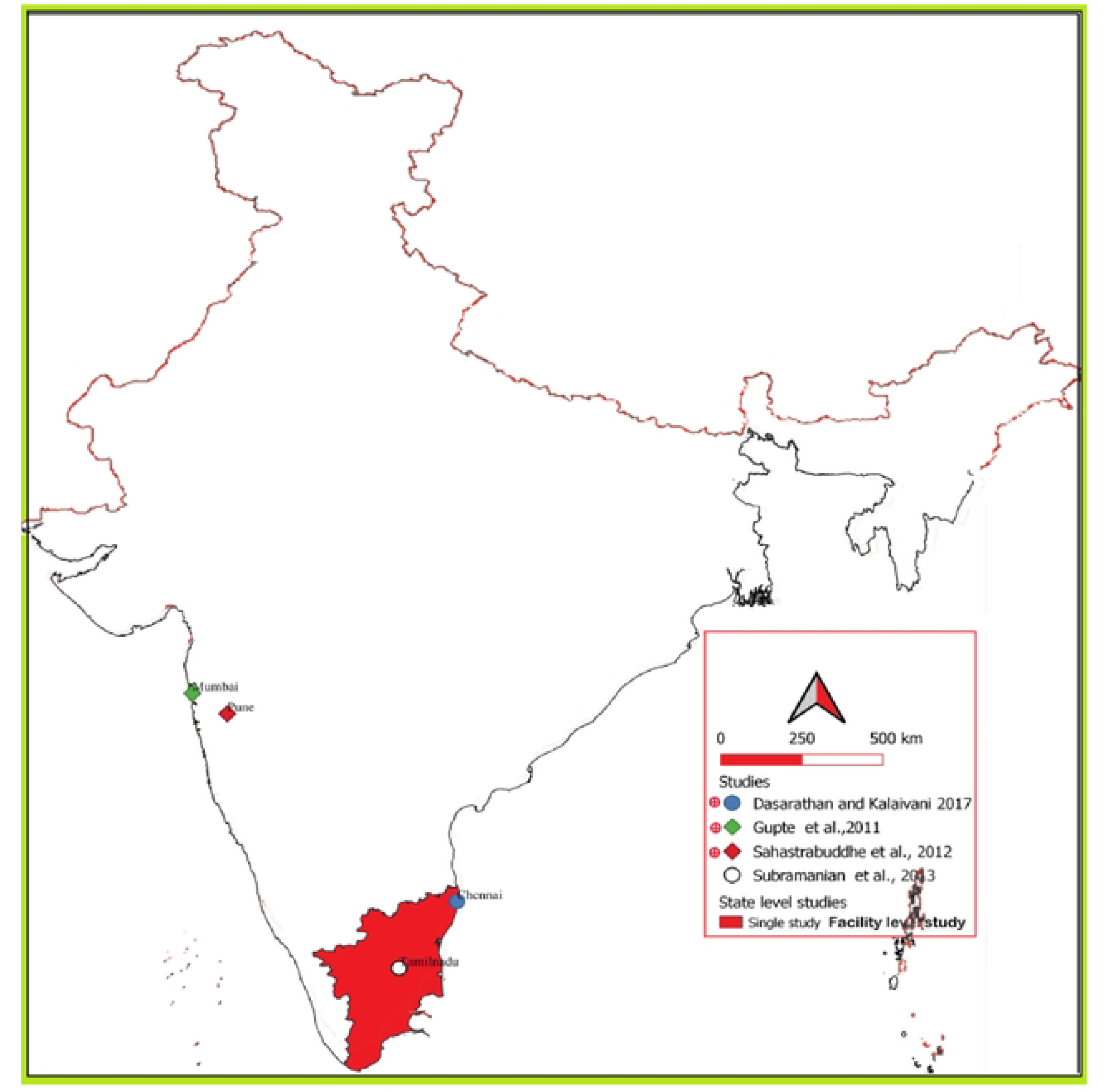
Geographical representation of published four curable STls among H/TGs in India.

### Findings among PWID

Prevalence of STI among PWID reported in four articles [11, 42–44]. Mahanta et al. [42] published a cross-sectional study on PWID in Manipur, Nagaland, Mumbai, and Thane. Syphilis seroprevalence was reported as 19.5% at Wokha, Nagaland. This was the highest reported syphilis prevalence in this study [Figure 6]. The highest reported prevalence (1.6%) of gonorrhoea was also reported in Wokha, and that of chlamydia (11.4%) was at Phek in Nagaland. This was published in 2008 but no specific study period was mentioned. An article comprised of district level data from North-eastern India viz. Phek, Wokha, Churachandpur, Bishnupur along with Mumbai-Thane region was also published. Prevalence data was reported on syphilis and chlamydia for PWID. Representation from North-eastern region was observed [Figure 6]. No STI prevalence data on PWID from West Bengal was found. The data also implied a weak regional representation of STI prevalence in India among PWID population [Table 6].

**Figure 6:**
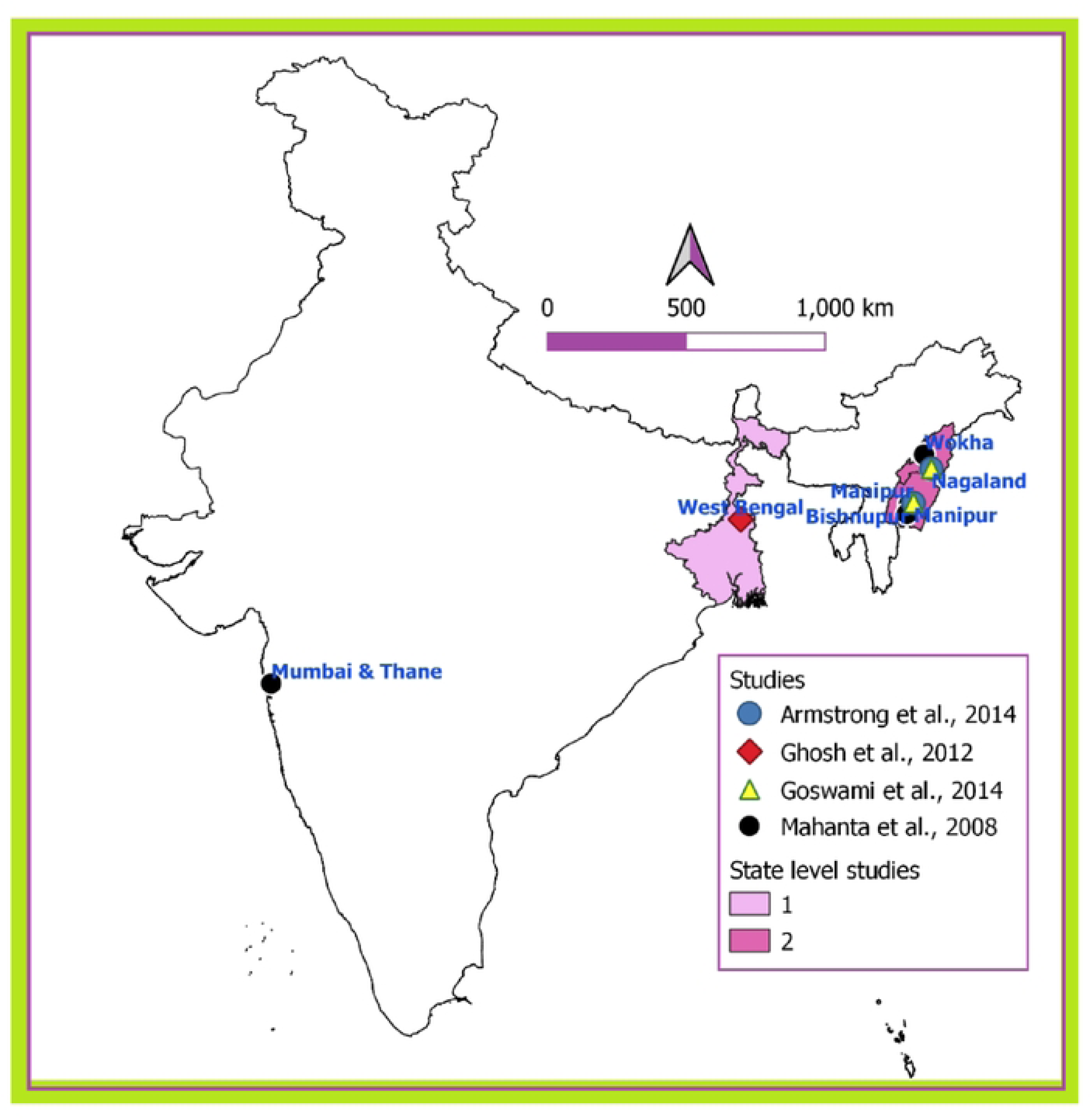
Representation of published articles on four curable STls among PWIDs in India.

### Quality of the selected studies

The modified Newcastle - Ottawa Quality Assessment Scale was applied to determined quality of included studies and the result was presented pictorially (Figure 7). Here, it must be noted that, quality of all the included studies calculated and presented here is only to accomplish the demands of present systematic review, except this authors have no purposeful intention (Figure 7).

**Figure 7:**
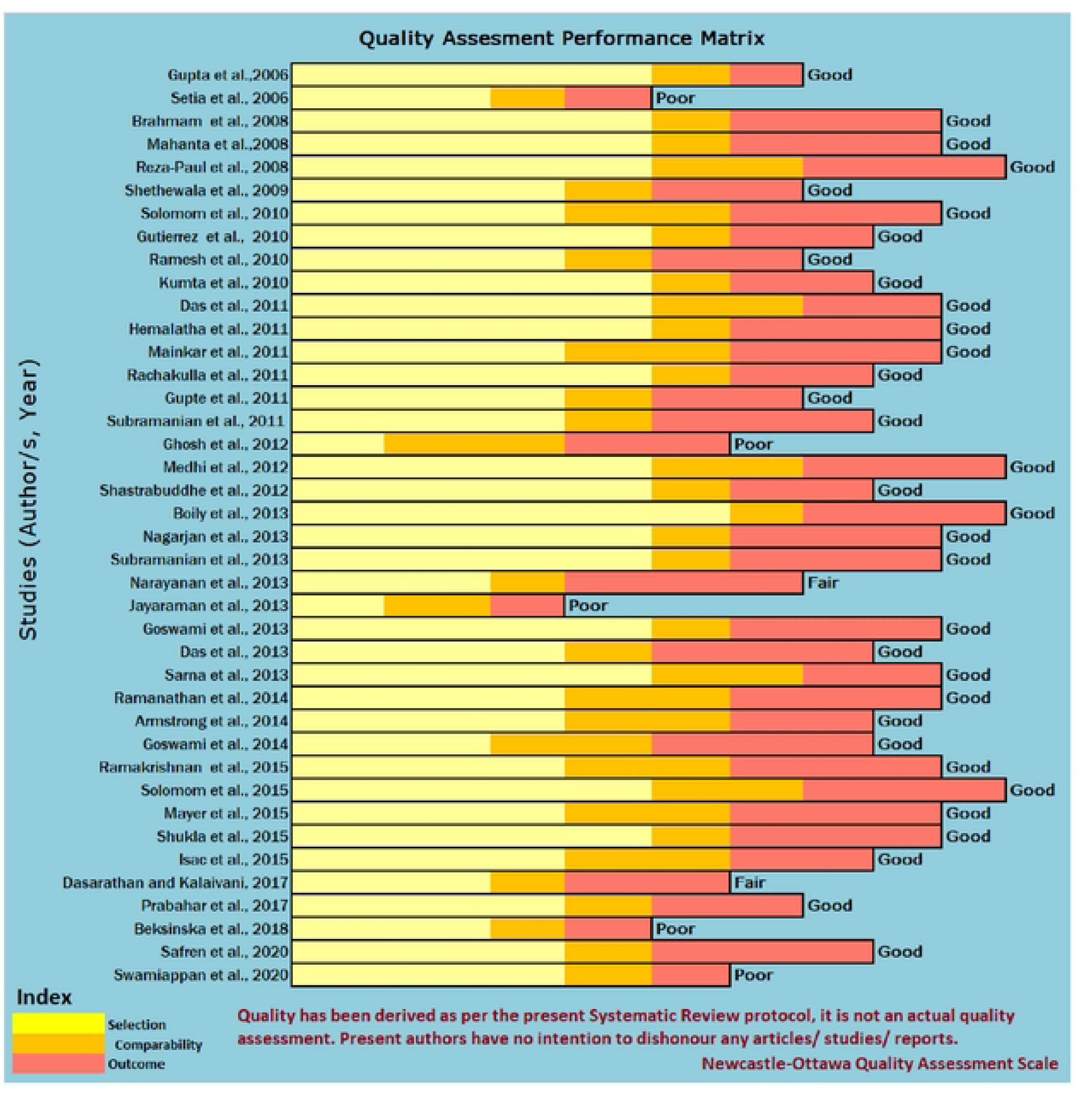
Quality Assessment Performance Matrix.

### Strength and limitations

The prevalence of STIs shows considerable heterogeneity by geographical setting and key population group. At the same time, limited numbers of study reported STI prevalence, sampling heterogeneity across studies prevent definite conclusions and how the prevalence of STIs varies among key populations. All the included studies in the present systematic review used probability-based sampling and both in facility as well as community-based setting. However, there were limited data points from all regions in India. The variation among included studies highlights a limitation of the present review, as the findings can vary based on the electronic databases and selected search terms. However, present article summarizes of all study that allows pre-defined acceptance criteria and qualify the quality assessment tool (Table 1-6).

## Discussion

The present systemic review had generated on the prevalence and spatiotemporal distribution of the four STIs in Indian key population. Prevalence of STIs showed extensive heterogeneity through spatiotemporal setting and people practising high-risk beheviour in India. A national STI surveillance cum prevention programme is essential among key populations in India. In the light of present findings and with the identified limitations, it can be concluded that existing HIV surveillance system under NACP, can be utilized with additional bio-specimen collection to determine STI prevalence among high risk populations. However, in future, with the availability of the comparable data from most of the regions in India, it will be possible to conduct a systemic review and meta-analysis on the spatiotemporal distribution of the four curable STIs in Indian general population.

## Data Availability Statement

No separate datasets were generated or analysed during the current study. All relevant data from this study are available with the present article.

## Supporting Information

PRISMA guideline (2020).

